# Back to the medical classes-Part I-Strategy for return to the presential practices during COVID-19 pandemics in a Brazilian Medical School

**DOI:** 10.1101/2021.10.14.21264918

**Authors:** Maria Auxiliadora Nogueira Saad, Vinicius César Jardim Pereira, Arnaldo Costa Bueno, Alan Araujo Vieira, Maria de Fátima Pombo March, Maria Dolores Salgado Quintans, Bruno Mendonça Barcellos, Laura da Cunha Ferreira, Karla Regina Oliveira Moura Ronchini, Luis Guillermo Coca Velarde, Claudete Aparecida Araújo Cardoso, André Ricardo Araujo da Silva

## Abstract

**Introduction:** In Brazil, practices of medical students have been interrupted due to COVID-19 to meet emergency demands.

**Aim:** To describe a strategy for a controlled return to the presential practices for medical students.

**Methods:** We developed a standardized protocol to be applied before and during the return of the practical classes in medical students of Universidade Federal Fluminense, in the follow months after COVID-19 pandemic beginning. The protocol was comprised in three parts: Remote training on COVID-19 prevention; Face-to-face training focused in COVID-19 prevention, handwashing and personal protective equipment use; Investigation of students COVID-19 status before starting practical activities and weekly monitoring for COVID-19 during seven weeks. The training was done by medical teachers in small groups for medical students of the last lective semester.

**Results:** The classes were interrupted on March 12, 2020 and returned in August 10, 2020. Seventy-one students were trained and followed. The mean age was 26.6 years (26.7 ±0.835) and 54% were female. Forty-nine (69%) students over 71 had a private health insurance, 60 (84.5%) shared a house/apartment with one or more person and 12(16.9%) reported a previous comorbidity. Eighteen (25.4%) over 71 reported previous symptoms of COVID-19, being positive in two students. During the follow-up, fourteen (19.7%) over 71 students were placed in quarantine due to signals/symptoms compatible with COVID-19 or contact with symptomatic case. Only two cases (2.8%) were confirmed and occurred in Brazilian epidemiological week 37.

**Conclusion:** The protocol was successful in minimizing COVID-19 acquisition during practical classes of medical students.

## Introduction

The COVID-19, a new disease caused by virus SARS-COV-2 and described in December 2019, rapidly spread around the world causing the most important pandemic of the current century. ^1, 2^The infection is transmitted person-to-person by droplets, contact with inanimate surfaces contaminated with the virus or aerosol in specific situations. ^2,3^

Aiming to mitigate the spread of disease, countries have adopted several non-pharmaceutical interventions (NPI), being the most common: mass and small gathering cancellation; closure of educational institutions; border restriction; individual movement restrictions; national lockdown; quarantine and personnel protective measure. ^4,5^

Brazil has one-hundred thirty-seven public Medical schools and most of them were closed in March 2020, with little differences regarding the date. The last two years of Medicine course are dedicated to practical activities with patients (internship) and even these practices were interrupted.

Medical services, in a context of pandemic, could not be interrupted, creating a challenge: how to return to the practical activities in a safe way to avoid or minimizing COVID-19 students contamination? In this context, our aim was to describe a strategy for a controlled return to the presential practices for medical students.

## Methods

### Setting

School of Medicine-Universidade Federal Fluminense

### Study population

Medical students from School of Medicine

### School of Medicine description

School of Medicine from Universidade Federal Fluminense (UFF) is a public institution of Niterói city, Rio de Janeiro State, Brazil. The School was created in 1926 and is organized in twelve lective semesters, being the last two years of the course dedicated exclusively to the practical activities (internship) with patients. The course admits 90 students each new lective semester.

### Inclusion criteria

Medical students from the last lective semester (12^th^)

### Exclusion criteria

None

### Effects of COVID pandemic on Medical student practical classes

From March 12, 2020 to August 10, 2020 all practical classes were interrupted due to COVID-19 pandemic, following local regulamentation. Just some remote lectures were available during this period.

### Protocol description

A standardized protocol was developed by Coordination of internship to allow return of medical students of the last lective semester to the practical classes. The approved protocol for the major administration of the university was planned and executed in three phases:

#### Phase 1- Preparation and organization

In this phase, seven medical professors were capacitated in COVID-19 prevention, specifically in handwashing techniques, correct use of personnel protective equipment, as well other NPIs

#### Phase 2- Execution and training

The second phase consists of two stages

Stage 1- Two-hour remote lecture of COVID-19 contents (prevention, handwashing techniques, correct use of personnel protective equipment (PPE), secure behavior during the hospital stay).

Stage 2- Two-hour presential training for the whole group of the students consisting of the same themes presented in stage 1. The students were divided in small groups (until 5 students), each one with one medical professor.

Stage 3- Immediately after the presential training, all students were inquired about COVID-19 antecedents, existence of previous health diseases, type of medical assistance used (public or private health insurance), and presence or not of cohabitans.

#### Phase 3- Active weekly surveillance of COVID-19 signals/symptoms

All students were advised to report immediately any symptoms of COVID-19 during the return to the practical activities and in case of positive feedback, were removed from 10 days counting since the date of beginning of the symptoms and tested for SARS-COV-2 if possible.

The students were followed during the seven weeks of internship (until October 4, 2020).These weeks were the period necessary to students complete the Medicine course.

### Epidemiological setting of COVID-19 during the study

The months from March to July 2020 represented the first wave of COVID-19 in Rio de Janeiro state, Brazil. According to the Rio de Janeiro State epidemiology, the peak of cases occurred in Brazilian epidemiological week 18.^6^During the whole period of the study, no COVID-19 vaccine was available in Brazil.

### Ethical approval

The study was approved by Ethics Committee of School of Medicine (Universidade Federal Fluminense) under number 4.610.403. The Project is registered in Brazilian Platform of Researches (Plataforma Brasil) under register CAAE: 44666621.2.0000.5243, dated from March 24, 2021. A consent statement for the use of data from the students were required for each person included.

## Data analysis

The data were collected in an Excel file and a descriptive analysis was conducted. Means with confidence interval were calculated for numeric variables. We used density of incidence (cases of COVID-19 suspected cases/1000 persons-days) to calculate possible new COVID-19 cases in medical students during the surveillance period. The occurence of suspected cases were classified according to the Brazilian epidemiological week

## Results

Seventy-one students of the last lective semester were trained in remote and practical classes and followed during seven weeks. The demographic data ofparticipants are shown inthetable 1.

**Table 1.** Demographic data of 12^th^–period Medical students (Universidade Federal Fluminense-August 2020)

| Variable | Values |
| --- | --- |
| **Age in mean** (95% confidence interval) | 26.6 (26.662 ±0.835) |
| **Gender (N)** |  |
| -Female (%) | 40 ( 54) |
| -Male ( %) | 31 (46) |
| **Health insurance (N)** |  |
| -Public (%) | 22 (31) |
| -Private (%) | 49 (69) |
| **Presence of cohabitans (N)** |  |
| -None (%) | 11 (15.5) |
| - One or more persons (%) | 60 (84.5) |
| **Comorbidity presence ( N)** |  |
| - Yes (%) | 12 ( 16.9) |
| - No ( %) | 59 (83.1) |

Twelve students declared presence of one or more comorbidity, being asthma reported by seven students, allergic rhinitis in two, atopic dermatitis in two and depression in one.

About specific investigation of COVID-19 antecedents occurred in students before the beginning of practical classes, eighteen students reported previous symptoms of disease and nine of them did specific tests to detected SARS-COV-2. Only 2 (11.1%) related positive tests for SARS-COV-2 over the 18 suspected cases.

During the surveillance period of students after beginning of practical classes, ten students (14%) were placed in quarantine due to signals/symptoms of COVID-19.Of symptomatic students, RT-PCR for SARS-COV-2 was negative in six (60%), positive in two (20%) and not performed in the two (20%) remaining. The suspected cases occurred in Brazilian Epidemiological week 33 (1 case), 35 (3 cases), 36 (two cases), 37 (three cases) and 39 (1 case).The confirmed two cases occurred in Brazilian Epidemiological week 37 in students that shared the same apartment. The density of incidence (confirmed cases/1000 persons-days) of confirmed cases were 0.57, representing an incidence of 2.8%. No admission or death of symptomatic medical students was reported during the follow-up period.

## Discussion

The Medical schools were also impacted with COVID-19 pandemics and most of them were closed as public health measures to mitigate new cases acquisition.^7,8^As consequence of Medical Schools closure, alternative strategies were created to maintain the classes emerging as online lectures, e-learning platforms, webcasting, virtual group discussions, mentoring and video-conferencing.^9^

The prolonged suspension of clinical practices could impact in the conclusion of Medicine curriculum and choice of future specialty, but surprisingly the adaptation and transformation on medical education caused by COVID-19 pandemics had generate positive expectations in students and professors. ^9,10^

A national lockdown in Brazil was not implemented, but regional legislation of different 26 States of the country regulated restrictive measures including School of Medicine closures. Following local regulation, all undergraduate activities of UFF, including Medical School were fully interrupted on March 12, 2020. Considering that practical activities were necessary to complete formation of last-period medical students, we proposed a standardized protocol to allow return of this specific stage and provide human resources to fight COVID-19.

The focus of our training was to provide knowledge of students about COVID-19 prevention and adequate use of PPE, in addition to safe behavior during hospital stay, since no standardized treatment or vaccine were available at that moment and lack of adequate training is an important gap during COVID-19 health assistance described by Brazilian healthcare workers. ^11^

One of our concerns was to recognize student’s accessibility to Brazilian health system, if necessary, in case of symptomatic disease. In Brazil, access to health system is free and guarantee for the whole population, despite a considerable percentage (around 54%) of population pay for a private health insurance.^12^ In our study population we verified a higher rate of utilization of private health insurance compared with reported by Castro and cols. ^12^

Other important topic included in our protocol was to identify number of persons sharing the same house/apartment. The higher percentage (84.5%) of students living with one or more persons could be an important information mainly to identify possible infected roommates and localized clusters of disease. A systematic review of SARS-COV-2 of secondary attack rate of COVID-19 in household contactsfound that rate varies widely across countries, being from 4.6% to 49.6%. ^13^

Monitoring suspected cases with laboratory diagnosis, if possible and quarantine are measures strongly recommended to mitigate the spread of COVID-19. ^14^ Both of them were included in our protocol and allow to identify fourteen suspected cases during the seven-week surveillance. The incidence (2.8%) of confirmed cases in suspected students in our study was lower than reported for healthcare workers where the incidence could reach until 21.5%. ^15-17^

At least two reasons could explain these values: the adherence to recommendation of our protocol and favorable epidemiological setting of the city during the study period (trends of reducing confirmed COVID-19 cases and deaths).

In the first months of pandemics there was a difficulty to perform diagnostic due to low offer and higher demand in the Rio de Janeiro, state. But even with this limitation of resources it was possible to investigate the disease in almost 80% of suspected cases.

Our study has some limitations. The first one is to be conducted during a period where the epidemiological setting of the state was favorable, which could interfere positively in results. The second one is the difficulty to confirm the infection in all suspected cases, which was a reality of Brazilian healthcare system, during the COVID-19 pandemics.

In conclusion, our protocol was successful in minimizing COVID-19 acquisition during practical classes of medical students of Universidade Federal Fluminense. A continuous surveillance is necessary to investigate adequacy of the protocol according to the less favorable epidemiological setting.

## Data Availability

All data produced in the present work are contained in the manuscript

## Acknowledgments

We thanks to all medical coordinators of internship for this research

